# Trade-offs in Cardiovascular Risk Prediction Using Race and Social Determinants of Health

**DOI:** 10.64898/2026.04.02.26350089

**Authors:** Noah Hammarlund, Xiangren Wang, David Grant, Duncan Purves

**Author notes:** Corresponding Author: Noah Hammarlund, PhD, PO Box 100195, Gainesville, FL, USA.

## Abstract

**Importance:** Health systems are increasingly adopting race-neutral cardiovascular risk prediction tools, yet no study has examined how these choices redistribute preventive treatment at the point of clinical decision-making, particularly for Black individuals who already bear a disproportionate cardiovascular burden.

**Objective:** To evaluate how including race, substituting social determinants of health (SDoH), or excluding both reshapes cardiovascular risk classification, calibration, fairness, and clinical decisions.

**Design:** Retrospective cohort study with repeated cross-validation and integrated decision-focused evaluation, using data from the Coronary Artery Risk Development in Young Adults (CARDIA) study, with baseline measures from 2010 and cardiovascular outcomes through 2021.

**Setting:** Community-based longitudinal cohort recruited across multiple U.S. cities.

**Participants:** 3,241 Black and White adults without known cardiovascular disease at baseline.

**Main Outcomes and Measures:** Three models predicting 10-year incident cardiovascular disease (CVD) were compared on predictive performance, calibration, group-based fairness metrics, and realized clinical utility quantifying overtreatment and undertreatment at the ACC/AHA 7.5% preventive treatment threshold. Models included clinical predictors plus race (Model 1), clinical predictors plus SDoH (Model 2), and clinical predictors only (Model 3).

**Results:** Among 3,241 participants (46% Black, mean age 50 years, 6.9% 10-year CVD incidence), overall predictive performance was similar across models (AUC 0.762 to 0.768). Predictor choice substantially reshaped clinical decisions at the guideline threshold. Model 2 improved parity metrics but produced systematic underprediction of risk and concentrated new overtreatment among Black participants. Model 3 further improved parity metrics but generated new undertreatment among Black participants, with four cases of untreated CVD and none avoided. No single evaluative dimension captured the full equity consequences of these design choices.

**Conclusions and Relevance:** Model design choices substantially reshaped treatment eligibility despite similar overall performance, with concentrated consequences among Black participants that are not apparent in population-average metrics. Parity metrics improved under both alternative models, yet both produced clinical harms that only became clear through holistic evaluation. The case for race removal has rested primarily on conceptual grounds, but comprehensive empirical evaluation is necessary before health systems can be confident their model choices truly serve the individuals most at risk.

## Introduction

In cardiovascular medicine, the American Heart Association recently introduced race-free risk equations as part of a broader shift away from race-inclusive predictive models.^1,2^ These changes are largely motivated by concerns that including race may reinforce inequities in care and normalize race-based clinical decision-making.^3–6^ Similar race-neutral transitions have occurred across clinical domains.^7–9^ An often proposed alternative is to replace race with social determinants of health (SDoH), which aim to capture the social and structural conditions that drive health disparities more directly and actionably.^10–13^

The consequences of these design choices are especially important in the cardiovascular context, where racial identification may capture important information about lived experiences and systemic disadvantage that shape cardiovascular risk.^14–16^ However, available SDoH measures may not fully reflect broader social and structural disadvantage, meaning neither race-inclusive nor race-neutral approaches are without limitation.^17,18^ Both the inclusion and exclusion of race may carry harms with implications for treatment allocation and access to preventive care for populations already facing disproportionate risk. Careful empirical evaluation is therefore needed to understand the practical consequences of these design choices. The case for race removal has rested primarily on conceptual grounds, with empirical evaluation focused narrowly on overall performance and calibration parity rather than the full distribution of clinical benefit and harm at the point of treatment.

This study provides such an evaluation in cardiovascular risk prediction, where guideline-based treatment thresholds translate predicted risk directly into clinical action. Using an integrated approach that jointly assesses predictive performance, calibration, group-based fairness, and realized clinical utility, we compare models that (1) include race, (2) replace race with detailed measures of SDoH, and (3) rely only on clinical predictors. These three models define two distinct comparisons: replacing race with SDoH, which tests whether social and structural context can substitute for racial identification, and removing race entirely, which tests the consequences of a strictly race-neutral approach. We assess how each design choice translates into preventive treatment eligibility at a guideline-recommended risk threshold, examining who gains or loses access to care and how clinical benefit and harm are distributed across racial groups.

## Methods

### Study Population

We used data from the Coronary Artery Risk Development in Young Adults (CARDIA) study^19^, a longitudinal cohort that recruited Black and White participants aged 18–30 years in 1985–86 from multiple U.S. cities, with representation across race and sex. CARDIA includes detailed longitudinal measures of clinical risk factors and SDoH, enabling direct comparison of race-based and SDoH-based prediction models. Of 5,113 participants enrolled at baseline, 1,707 were excluded due to missing Year 25 discrimination module data, 49 were excluded due to missing or unclassifiable income data, and an additional 116 were excluded due to missing binary clinical or social determinants of health variables, yielding a final analytic sample of 3,241 Black and White adults. This study was determined to be non-human subjects research by the University of Florida Institutional Review Board (IRB).

We used baseline data from 2010 (Year 25) to predict cardiovascular outcomes measured in Year 35 of the study (2021), which corresponds to a follow-up period of approximately 10-years. The primary outcome was incident cardiovascular disease (CVD), defined as myocardial infarction, stroke, heart failure hospitalization, or coronary revascularization. The CARDIA study identified morbidity endpoints from participant-reported hospitalizations and verified through medical-record abstraction and adjudication.

We modeled 10-year incident CVD as a binary outcome to align with risk-threshold–based clinical decisions and to aid evaluation of performance, fairness, and utility metrics. Predicted risk was translated into treatment recommendations using the 7.5% threshold from ACC/AHA cholesterol management guidelines, which reflects standard preventive care practice, most commonly for statin therapy but also as a broader trigger for intensified preventive management.^20–22^ We focused on the 7.5% threshold because it represents a discrete clinical action point at which prediction errors translate directly into treatment.

This study was reported in accordance with the Strengthening the Reporting of Observational Studies in Epidemiology (STROBE) guidelines for cohort studies.

### Predictors and Measures

Predictors included age, sex, race, blood pressure, lipid levels, body mass index, diabetes status, hypertension history, and kidney function, selected to align with major cardiovascular risk models such as the Pooled Cohort Equations, Framingham Risk Score, and PREVENT framework.^1,2,23,24^ Race was measured by participant self-identification and was interpreted primarily as a social classification reflecting lived experience and structural exposure, without making assumptions about underlying mechanisms. Kidney function was estimated using the race-neutral 2021 CKD-EPI equation, consistent with current clinical recommendations and to avoid introducing race adjustment through other predictors.^25^ CARDIA captured a particularly rich set of SDoH measures including socioeconomic status, healthcare access, housing stability, social support, employment, insurance coverage, financial strain, food insecurity, and experiences of discrimination. SDoH variables were operationalized as binary indicators reflecting the presence of each social exposure.

Continuous clinical variables (blood pressure, BMI, eGFR, LDL, and HDL) were imputed using the sample median where missing. All other variables were handled via complete-case analysis.

### Model Development

We developed three prediction models differing by predictor sets: a clinical model including race (Model 1), a clinical model substituting detailed SDoH for race (Model 2), and a clinical-only model excluding both race and SDoH (Model 3).

We trained models using LASSO-regularized logistic regression with 10-fold cross-validation. Given the relatively low prevalence of incident CVD (6.9%), we applied upsampling within training folds to address class imbalance. We selected the regularization parameter by minimizing log loss. We selected log loss as the primary tuning metric because it penalizes miscalibration across the full probability range and prioritizes well-calibrated risk estimates, which are essential for threshold-based fairness and realized-utility analyses.^26–30^ We selected LASSO-regularized logistic regression over gradient-boosted tree models, which offered only marginal performance gains at the cost of interpretability.

### Statistical Comparisons of Model Performance and Fairness Metrics

Model comparisons used subject-level pooled-null permutation testing with 5,000 resamples across repeated cross-validation to account for correlated evaluations.^31–33^ Differences were assessed for AUC, sensitivity, specificity, and false-positive and false-negative rates. Sensitivity reflects the proportion of CVD cases correctly classified as high-risk. Specificity reflects the proportion of non-cases correctly classified as low risk. We also assessed calibration, which measures whether predicted probabilities are systematically too high or too low and whether they are appropriately spread, overall and stratified by race.^34^

Additionally, we compared seven group-based fairness metrics (demographic parity, equal opportunity, equalized odds, disparate impact, predictive parity, average odds, and false-positive rate difference) each of which quantifies a different aspect of how classification errors and treatment rates are distributed between Black and White participants. Supplemental Table 1 provides formal definitions of each metric. All fairness metrics evaluated in this study assess group-level, relative differences in classification and error rates between racial groups, rather than individual-level notions of fairness.

McNemar’s tests evaluated whether paired changes in treatment classification reflected systematic directional shifts rather than random disagreement between models.^35,36^

### Utility Framework

To link model performance to treatment consequences near the clinical decision threshold, we adapted the utility framework of Coots et al. to evaluate realized rather than expected utility.^37^ Whereas Coots et al. evaluated expected utility under outcome uncertainty, we assessed realized utility using observed outcomes, allowing direct estimation of the gains and losses each model would have produced without assuming perfect calibration or treating predicted probabilities as true risk.

We quantified the clinical consequences of each model by assigning utility weights to treatment outcomes at the 7.5% threshold. Treatment outcomes were weighted as benefits, costs, or harms accordingly. Weights were calibrated so that the net benefit of treatment equals zero exactly at the guideline threshold, consistent with the benefit-harm trade-off that threshold encodes. Full parameterization is provided in the Supplemental Appendix.

Sensitivity analyses using alternative treatment thresholds (5% and 10%) were conducted to assess the robustness of results to threshold choice.

## Results

Table 1 summarizes demographic, clinical, and social characteristics of the analytic sample (N = 3,241). Participants were 46% Black and 54% White with a mean baseline age of 50 years. Black participants had higher prevalence of hypertension, diabetes, obesity, and antihypertensive medication use, as well as higher blood pressure and lower measured kidney function. Black participants also reported higher levels of financial strain, food insecurity, difficulty paying for medical care, lack of insurance coverage, and more frequent experiences of racial, socioeconomic, and gender discrimination.

**Table 1:** Descriptive characteristics of the study population by race. Baseline demographic, clinical, social, and outcome characteristics of participants, stratified by self-reported race. Continuous variables are reported as sample means. All other variables are reported as percentages representing the proportion of individuals in each racial group with the specified characteristic, with parenthetical definitions provided where applicable.

| Category | Variable | Overall | Black | White |
| --- | --- | --- | --- | --- |
| **Demographic** | N (sample size) | 3,241 | 1,495 | 1,746 |
|  | Age at Y25 (years) | 50 | 49.4 | 50.5 |
|  | Female (%) | 55.9 | 59.7 | 52.7 |
|  | Not in relationship (%) | 42.7 | 40.7 | 44.4 |
|  | Black race (%) | 46.1 | 100 | 0 |
| **Outcome** | Incident CVD (10-year) (%) | 6.9 | 9.1 | 5 |
| **Clinical** | BMI (kg/m<sup>2</sup>) | 29.9 | 31.9 | 28.2 |
|  | Obese (BMI ≥30) (%) | 43.3 | 55.9 | 32.4 |
|  | Systolic BP (mmHg) | 118.6 | 123.2 | 114.6 |
|  | Systolic BP ≥130 mmHg (%) | 39.9 | 51.6 | 29.9 |
|  | Diastolic BP (mmHg) | 73.9 | 77.3 | 70.9 |
|  | Diastolic BP ≥80 mmHg (%) | 25.1 | 35.5 | 16.3 |
|  | HDL (mg/dL) | 58 | 57.5 | 58.5 |
|  | Low HDL (%) | 13.7 | 12.4 | 14.9 |
|  | LDL (mg/dL) | 112.1 | 111.3 | 112.7 |
|  | LDL ≥160 mg/dL (%) | 7.6 | 8.2 | 7.1 |
|  | eGFR (mL/min/1.73 m<sup>2</sup>) | 92.9 | 90.1 | 95.2 |
|  | eGFR ≤60 (%) | 2.4 | 4.1 | 0.9 |
|  | History of diabetes (%) | 10.7 | 14.8 | 7.2 |
|  | History of hypertension (%) | 32.7 | 45.9 | 21.4 |
|  | On antihypertensive medication (%) | 26.9 | 38.5 | 16.9 |
|  | On statin (%) | 15.5 | 16.7 | 14.4 |
|  | Current smoker (%) | 21.7 | 16.7 | 26.1 |
| **Social**<br>**Determinants**<br>**of Health** | Low education (≤ high school) (%) | 40.4 | 53.6 | 29 |
|  | Low income (lowest category) (%) | 17.2 | 27.4 | 8.5 |
|  | Currently employed (%) | 79.8 | 73.5 | 85.2 |
|  | Financial strain (%) | 57.3 | 61.6 | 53.6 |
|  | Food insecurity (%) | 15.8 | 22.5 | 10 |
|  | Housing instability (%) | 1.5 | 1.7 | 1.3 |
|  | Healthcare access barrier (%) | 11.1 | 15.1 | 7.6 |
|  | Uninsured (%) | 27 | 34.3 | 20.7 |
|  | Public insurance (%) | 11 | 18.2 | 4.8 |
|  | Trouble paying for food/heating (%) | 27.9 | 37.1 | 20.1 |
|  | Trouble paying for medical care (%) | 32.6 | 37.9 | 28 |
|  | Any close friend or family member (%) | 99.4 | 99.3 | 99.5 |
| **Discrimination**<br>**&**<br>**Psychosocial** | Racial discrimination (any) (%) | 24.7 | 46.1 | 6.4 |
|  | SES discrimination (any) (%) | 16.8 | 26.4 | 8.5 |
|  | Gender discrimination (any) (%) | 26.5 | 38.1 | 16.7 |
|  | Weight discrimination (any) (%) | 10.6 | 12.6 | 8.9 |

### Overall Model Performance

Overall predictive performance was similar across all three models, with AUC ranging from 0.762 to 0.768 and no statistically significant differences (Table 2). Substituting SDoH for race shifted the sensitivity–specificity trade-off without materially changing overall performance. Model 2 identified more CVD cases overall but also referred more individuals who would not develop CVD for preventive treatment. The clinical-only model performed comparably to the race-inclusive model across all metrics. Predicted probabilities were appropriately spread across the risk range in all three models, but Model 2 showed systematic underprediction of absolute risk overall and among both racial subgroups, while Models 1 and 3 were well-calibrated overall, though Model 3 showed modest overprediction among Black participants (Supplemental Table 2).

**Table 2.** Overall model performance metrics. Summary of overall model performance metrics for three predictive models: Model 1 (Clinical + Race), Model 2 (Clinical + Social Determinants of Health), and Model 3 (Clinical Only). Metrics include the area under the receiver operating characteristic curve (AUC), log-loss, sensitivity, specificity, false-positive rate (FPR), and false-negative rate (FNR). Values represent averages across repeated cross-validation with permutation testing. Asterisks (*) denote statistically significant differences relative to Model 1 (p < 0.05). Model 2 achieved slightly higher sensitivity but lower specificity, while Model 3 showed the most conservative classification pattern with comparable overall performance.

| Metric | Model 1 (Race) | Model 2 (SDoH) | Model 3 (Trad.) |
| --- | --- | --- | --- |
| AUC | 0.768 | 0.762 | 0.768 |
| Log Loss | 0.219 | 0.224 | 0.219 |
| Sensitivity | 0.642 | 0.671 * | 0.641 |
| Specificity | 0.761 | 0.720 * | 0.763 |
| FPR | 0.239 | 0.280 * | 0.237 |
| FNR | 0.358 | 0.329 * | 0.359 |

### Differences by Race

Table 3 shows race-stratified results. Overall performance was similar across groups, but the pattern of errors differed meaningfully by model and race. Among Black participants, substituting SDoH for race produced minimal change in sensitivity but increased false positives. Model 2 referred more Black individuals who would not develop CVD than Model 1. Removing race entirely had the opposite effect. Model 3 reduced false positives among Black participants but slightly reduced sensitivity, meaning fewer true CVD cases were identified. Among White participants, the SDoH-based model substantially increased sensitivity at the cost of more false positives, a pattern that was partly reversed under the clinical-only model. Gains in sensitivity for one group were consistently accompanied by losses for the other. Reclassification testing confirmed these shifts were systematic rather than random.

**Table 3.** Differences in model performance and fairness metrics stratified by racial group. Model performance metrics stratified by racial group subsample. Values are shown separately for Black and White participants for each model: Model 1 (Clinical + Race), Model 2 (Clinical + Social Determinants of Health), and Model 3 (Clinical Only). Metrics include the area under the receiver operating characteristic curve (AUC), log-loss, sensitivity, specificity, false-positive rate (FPR), and false-negative rate (FNR), averaged across repeated cross-validation with permutation testing. Asterisks (*) indicate statistically significant differences relative to Model 1 within each racial subgroup (p < 0.05).

| Metric | Model 1 (Race) | Model 2 (SDoH) | Model 3 (Trad.) |
| --- | --- | --- | --- |
| <u>Black Subsample</u> |  |  |  |
| AUC | 0.754 | 0.746 | 0.756 |
| Log Loss | 0.268 | 0.273 | 0.268 |
| Sensitivity | 0.725 | 0.728 | 0.710 |
| Specificity | 0.658 | 0.630 * | 0.681 * |
| FPR | 0.342 | 0.370 * | 0.319 * |
| FNR | 0.275 | 0.272 | 0.290 |
| <u>White Subsample</u> |  |  |  |
| AUC | 0.763 | 0.764 | 0.763 |
| Log Loss | 0.178 | 0.181 | 0.178 |
| Sensitivity | 0.514 | 0.581 * | 0.533 |
| Specificity | 0.846 | 0.794 * | 0.829 * |
| FPR | 0.154 | 0.206 * | 0.171 * |
| FNR | 0.486 | 0.419 * | 0.467 |

### Fairness Metrics

Parity-based fairness metrics generally improved as race was removed or replaced, but the pattern was partly mechanical. Excluding race constrains the model from stratifying by group, which narrows measured gaps by construction rather than by improving equitable performance. Results are shown in Figure 1 and Supplemental Table 3.

**Figure 1.**
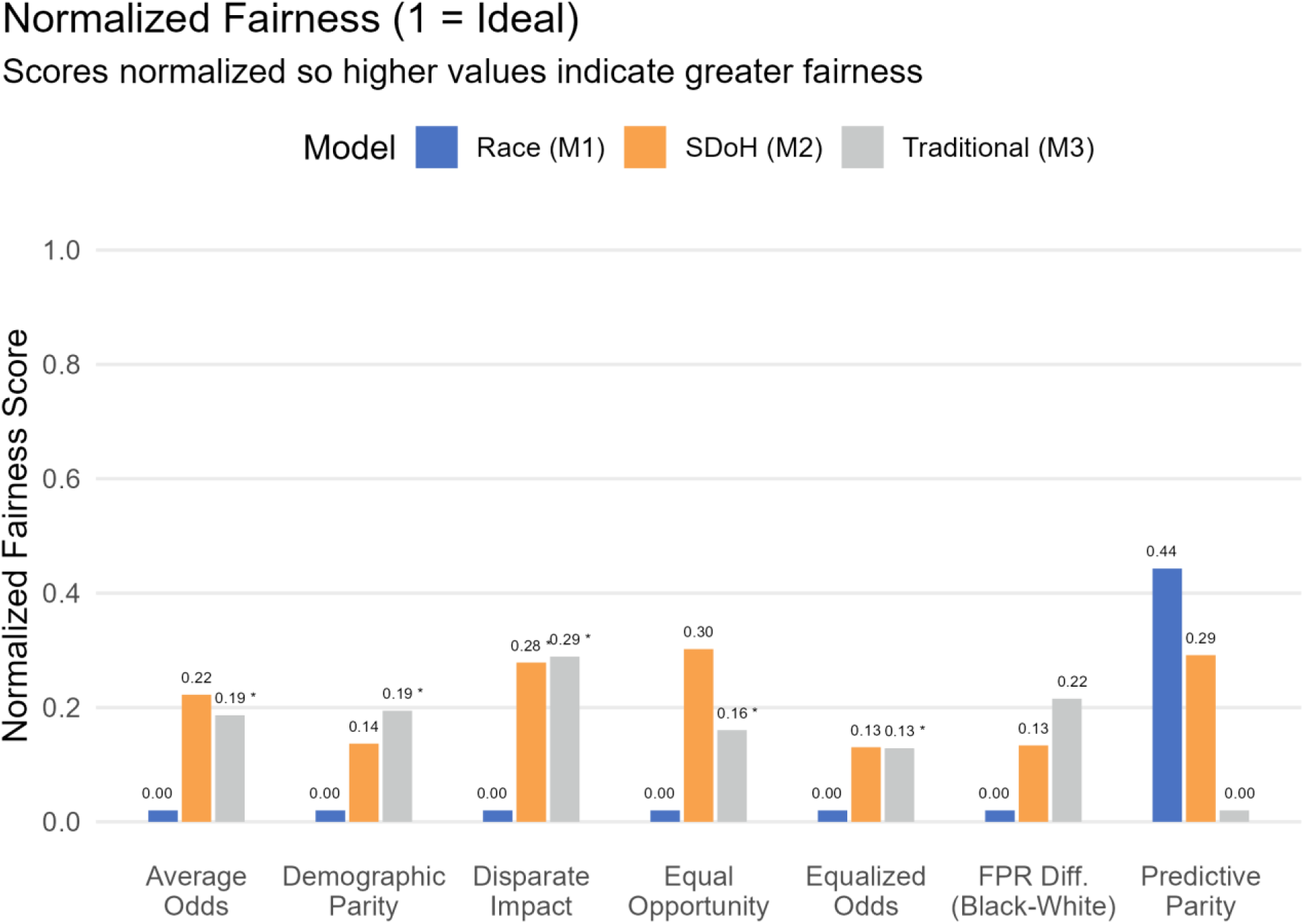
Fairness Metrics Across Models. Parity- and error-based fairness metrics for each model, including demographic parity, disparate impact, equal opportunity, average odds, equalized odds, false-positive rate difference (Black–White), and predictive parity. For all ratio-based metrics, values closer to one indicate smaller between-group disparities (parity), while greater deviation from one reflects larger inequities. Models excluding race reduced several parity gaps primarily through shifts in classification rates rather than improved accuracy.

Model 2 reduced several between-group disparities relative to Model 1, with the most notable improvement in disparate impact, which moved meaningfully closer to parity. Model 3 produced the largest reductions in parity gaps overall, with significant improvements across most fairness metrics. No model consistently minimized disparities across all fairness definitions, and residual differences persisted under every model. Sensitivity analyses at 5% and 10% risk thresholds yielded consistent results, with the direction and relative ordering of fairness metrics stable across all thresholds.

### Utility

Figure 2 shows the realized clinical consequences of reclassification at the 7.5% treatment threshold, the point at which predicted risk triggers a recommendation for statin therapy or other preventive intervention, decomposed into overtreatment and undertreatment gains and losses by racial group. Population-average utility differences across models were small, but this aggregate picture masked concentrated subgroup effects among individuals near the treatment boundary.

**Figure 2.**
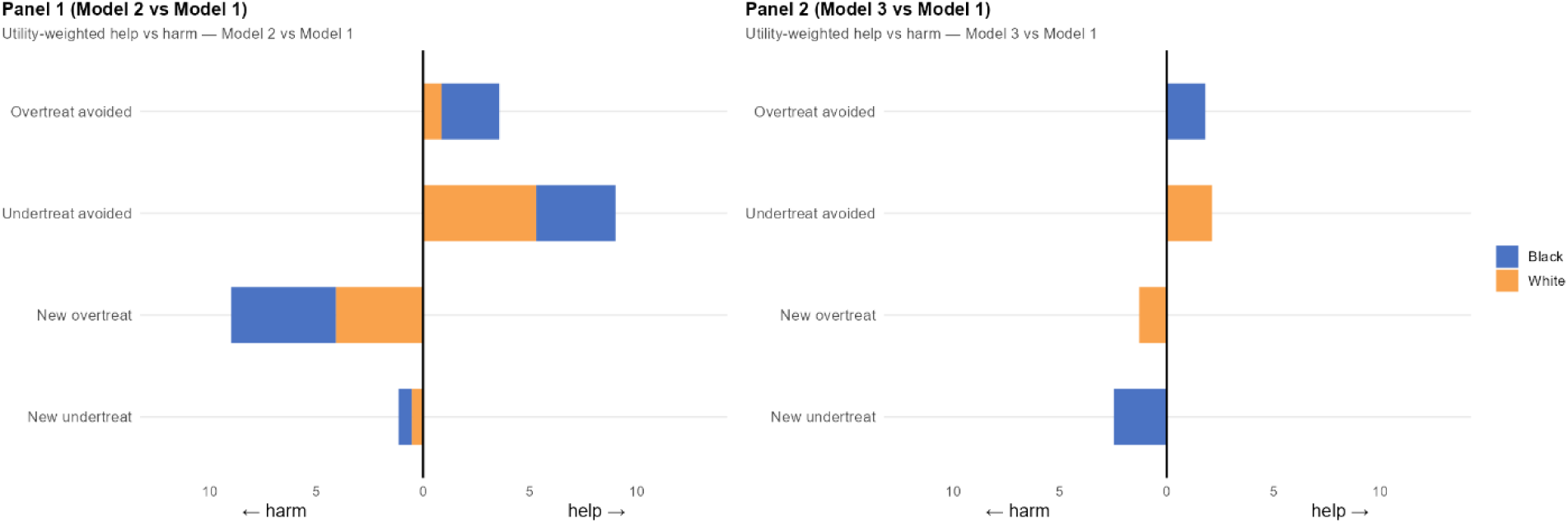
Clinical Reclassification / Utility Consequences. Stacked bars show realized utility associated with patient reclassification between models at the 7.5% treatment threshold. Reclassification is decomposed into four components: new overtreatment, overtreatment avoided, new undertreatment, and undertreatment avoided. Bar width represents the magnitude of clinical impact in each category, weighted by realized utility. Blue segments reflect impacts among Black participants and orange segments reflect impacts among White participants, with segment widths indicating each group’s contribution to the total effect. Arrows indicate whether reclassification represents clinical benefit or harm. Results are shown for comparisons between the SDoH-based and race-based models (left) and between the clinical-only and race-based models (right).

Substituting SDoH for race (Model 2 vs Model 1) produced the most reclassification, affecting 8.8% of individuals overall and 10.6% of Black participants. Among those reclassified, the dominant effect was new overtreatment. Most newly reclassified individuals were referred for preventive treatment but would not have developed CVD. Among Black participants, Model 2 avoided more undertreatments than it created, producing a modest net benefit in undertreatment reduction. However, this came at the cost of a substantial increase in overtreatment, concentrated largely among Black individuals.

Removing race entirely (Model 3 vs Model 1) produced fewer reclassifications overall, but among Black participants, the reclassifications that did occur were concentrated in undertreatment rather than overtreatment. Among Black participants, Model 3 generated four new cases of untreated CVD with no offsetting reduction in undertreatment elsewhere in the group. Nearly all reclassifications under both comparisons occurred within one percentage point of the 7.5% threshold, reflecting how small shifts in predicted risk near the treatment boundary drive meaningful changes in who receives care. Sensitivity analyses at alternative thresholds produced consistent directional findings.

### Predictor Importance

Across all three models, traditional clinical risk factors, particularly kidney function, blood pressure, lipid levels, diabetes, and hypertension history, accounted for the majority of predictive power (Supplemental Table 4). Including race or substituting detailed SDoH measures did not meaningfully alter the ranking of core clinical predictors.

## Discussion

This study examined how alternative modeling choices about race and SDoH reshape risk classification and preventive treatment decisions at a real-world clinical threshold. Across all three models, overall predictive performance was similar, yet gains in sensitivity for one racial group were accompanied by losses for the other. No model improved identification of high-risk individuals across both groups simultaneously. Parity metrics improved as race was removed or replaced, but this pattern was partly mechanical: excluding race constrains the model from stratifying by group, which narrows gaps by construction rather than by improving equitable performance. Calibration worsened under SDoH substitution, meaning apparent equity gains may partly reflect compressed risk estimates rather than genuinely more equitable classification. Realized utility analysis also revealed concentrated subgroup harms that were invisible in population-average metrics. Together, these results demonstrate that no single evaluative dimension captures the full consequence of these choices. Empirical evaluation across all dimensions is necessary to make the trade-offs visible to support deliberate clinical decision-making.

Predictor choice fundamentally shapes what a model represents and how its outputs can inform clinical action. Including race may better capture cumulative structural disadvantage that improves identification of Black individuals at high risk, but it does so by encoding group membership as a direct input to treatment decisions. Across both comparisons, that trade-off was visible among Black individuals, where neither alternative model matched the sensitivity achieved under Model 1. Residual disparities persisted under every model, reflecting how correlated clinical predictors continued to stratify risk even without race as an explicit input.

For the substitution comparison, SDoH substitution reflects a fundamentally different clinical logic than race removal. Unlike race, SDoH correspond to specific social conditions that may be directly targeted by intervention, even if they capture a narrower portion of contextual risk. Substituting race with SDoH therefore reflects a deliberate shift in what aspects of risk are emphasized rather than a simple loss of predictive fidelity. Because prediction models estimate risk rather than causal effects, changes in predictor sets redistribute how contextual factors influence risk estimates, with direct consequences for who crosses the treatment threshold.

Excluding race modestly reduced sensitivity for Black patients and increased undertreatment risk. Among Black participants specifically, the clinical-only model produced four new cases of untreated CVD, each representing an individual who would not have qualified for preventive treatment at the guideline threshold despite eventually developing disease. Notably, even with CARDIA’s unusually rich and longitudinal social data, replacing race did not fully recover sensitivity among Black participants. In typical clinical settings, where SDoH data are often sparse or inconsistently documented, these trade-offs may be more pronounced.

As health systems implement race-neutral cardiovascular risk tools, including PREVENT-based calculators, the primary justification for race removal has been conceptual. Empirical evaluation in that work was largely secondary, focused on confirming that calibration was similar across racial groups rather than examining how predictor choices redistribute clinical benefit and harm at the point of treatment. Our findings show that conceptual reasoning and calibration parity together are insufficient to characterize the full equity consequences of these design choices. A holistic empirical picture integrating performance, calibration, fairness, and realized clinical outcomes is necessary for responsible adoption decisions. Without it, clinicians, patients, and communities bearing disproportionate cardiovascular burden cannot meaningfully deliberate about which trade-offs are acceptable.

## Limitations

This study has several limitations. First, the analytic sample was restricted to participants identified as Black or White, while race was operationalized as a binary variable, which does not capture the full diversity of racial and ethnic identities or the heterogeneity of lived experiences that shape cardiovascular risk.

Second, SDoH were treated as static predictors, although social conditions such as employment, housing, and insurance coverage evolve over time. Models incorporating time-varying social exposures may better represent contextual risk.

Third, despite robust resampling and statistical comparisons, findings remain specific to the CARDIA cohort and may differ in populations with different baseline risks or distributions of social disadvantage. However, the evaluation framework itself is broadly applicable across clinical settings.

Fourth, the utility function was anchored to the guideline treatment threshold, reflecting standard clinical decision rules rather than individual patient preferences or heterogeneous treatment benefit. However, sensitivity analyses using alternative thresholds yielded similar patterns, supporting the robustness of the findings to this specification.

Finally, the analytic sample was restricted to participants with complete Year 25 discrimination module data. Prior CARDIA research has demonstrated that participants missing this module were disproportionately Black and socioeconomically disadvantaged.^38^ Risk and fairness estimates among Black participants may therefore be conservative.

## Conclusion

Substituting SDoH for race in cardiovascular risk prediction produced similar overall accuracy but systematically reshaped treatment eligibility near clinical decision thresholds. These shifts produced concentrated subgroup consequences that were not visible in population-average metrics alone. Excluding race modestly reduced sensitivity for Black individuals and increased undertreatment risk, while race-inclusive models identified more high-risk individuals at the cost of higher false positives. Even with unusually detailed social data, SDoH did not fully capture the contextual inequities that race often proxies.

No single model simultaneously optimized accuracy, calibration, parity, and clinical outcomes, and no single evaluative dimension was sufficient to reveal that. The conceptual case for race removal is important, but it requires a fuller empirical foundation before adoption decisions can be responsibly made. Integrated evaluation across all four dimensions provides that foundation. Clinicians, patients, and health-system leaders deserve that evidence before these decisions are made on their behalf.

## Acknowledgments

This manuscript was prepared using CARDIA Research Materials obtained from the NHLBI Biologic Specimen and Data Repository Information Coordinating Center and does not necessarily reflect the opinions or views of the CARDIA or the NHLBI. The authors thank Serena Jingchuan Guo and Jiang Bian for their early contributions to the conceptual development of this work. ChatGPT (GPT-4 and GPT-5, OpenAI, November 2024-December 2025) and Claude (Sonnet 4.6, Anthropic, January-March 2026) were used for editorial support, code refinement, code annotation, and iterative discussion during manuscript development. These tools did not contribute to the scientific design of the study, the analytic strategy, or the interpretation of results. All substantive decisions and responsibility for the work are the authors’ own. N.H. had full access to all the data in the study and takes responsibility for the integrity of the data and the accuracy of the data analysis.

## Data Availability

The CARDIA data used in this study are available through the NHLBI Biologic Specimen and Data Repository Information Coordinating Center (BioLINCC). Investigators may request access by submitting a research proposal and data use agreement through the BioLINCC website (https://biolincc.nhlbi.nih.gov/). The authors do not have permission to share the individual-level data directly.

## Author Contributions Statement

N.H.: Conceptualization, Methodology, Formal Analysis, Validation, Visualization, Writing – Original Draft, Writing – Review & Editing, Project Administration, Supervision. X.W.: Data Curation, Software, Formal Analysis, Writing – Review & Editing. D.G.: Conceptualization, Methodology, Writing – Review & Editing. D.P.: Conceptualization, Writing – Review & Editing.

